# Causal Evaluation of Laboratory Markers in Type 2 Diabetes on Cancer and Vascular Diseases Using Various Mendelian Randomization Tools

**DOI:** 10.1101/2020.08.21.20179622

**Authors:** Heejin Jin, Sanghun Lee, Sungho Won

## Abstract

Multiple studies have demonstrated the effects of type 2 diabetes (T2D) on various human diseases; however, most of these were observational epidemiological studies that suffered from many potential biases including reported confounding and reverse causations. In this article, we investigated whether cancer and vascular disease can be affected by T2D-related traits, including fasting plasma glucose (FPG), 2-h postprandial plasma glucose (2h-PG), and glycated hemoglobin A1c (HbA1c) levels, by using Mendelian randomization (MR). The summary statistics for FPG, 2h-PG, and HbA1c were obtained through meta-analyses of large-scale genome-wide association studies that included data from 133,010 non-diabetic individuals from collaborating Meta-Analysis of Glucose and Insulin related traits Consortium studies. Thereafter, based on the statistical assumptions for MR analyses, the most reliable approaches including inverse-variance-weighted (IVW), MR-Egger, MR-Egger with a simulation extrapolation (SIMEX), weighted median and MR-Pleiotropy RESidual Sum and Outlier (MR-PRESSO) methods were applied to identify traits affected by FPG, 2h-PG, and HbA1c. We found that coronary artery disease is affected by FPG, as per the IVW [log odds ratio (logOR): 0.21; *P*=0.012], MR-Egger (SIMEX) (logOR: 0.22; *P*=0.014), MR-PRESSO (logOR: 0.18; *P*=0.045), and weighted median (logOR: 0.29; *P*<0.001) methods, but not as per the MR-Egger (logOR: 0.13; *P*=0.426) approach. Furthermore, low-density lipoprotein cholesterol levels are affected by HbA1c, as per the IVW (beta (B): 0.23; *P*=0.015), MR-Egger (B: 0.45; *P*=0.046), MR-Egger (SIMEX) (B: 0.27; *P*=0.007), MR-PRESSO (B; 0.14; *P*=0.010), and the weighted median (B: 0.15; *P*=0.012) methods. Further studies of the associated biological mechanisms are required to validate and understand the disease-specific differences identified in the TD2-related causal effects of each trait.

## 1 Introduction

Type 2 diabetes (T2D) is characterized by high blood sugar, insulin resistance, and a relative lack of insulin and represents a common metabolic disorder worldwide. In its early stage, T2D is easy to ignore due to the lack of symptoms; however, chronic or poorly controlled T2D leads to eventually disabling or life-threatening complications. Numerous epidemiological studies have consistently demonstrated increased risks of cancer, vascular disease, nerve damage, and poor health-related outcomes in T2D patients (De Vegt et al., 1999;Laakso, 1999;Tsilidis et al., 2015), resulting in a shorter life expectancy (Collaboration, 2011). The main T2D-related complications reported in large-scale epidemiological studies tend to be malignant solid tumors (Johnson et al., 2012) and cardiovascular disease, including ischemic heart disease and stroke (Nesto, 2001;Bax et al., 2007;Gleissner et al., 2007;Young et al., 2009). However, the causal relationship between T2D and diverse health-related outcomes needs to be investigated and compared.

Fasting plasma glucose (FPG) levels ≥126 mg/dL or post-challenge 2-h plasma glucose (2h-PG) levels ≥200 mg/dL in a 75-g 2-h oral glucose tolerance test (2h-OGTT) have been used as diagnostic criteria for T2D. Additionally, hemoglobin A1c (HbA1c) levels ≥6.5% were added to these diagnostic criteria in 2010 (Gavin III et al., 1997;Association, 2010). The three tests (FPG, 2h-PG, and HbA1c) are dependent on blood glucose metabolism status. Specifically, FPG assesses the state of stable sugar levels in the body following a temporary increase in externally administered sugar. The 2h-OGTT indicates how efficiently insulin is processed during metabolism in response to increased externally administered glucose. HbA1c reflects the average blood sugar level until immediately before the test and not at the time of sample collection, because hemoglobin increases with time and according to glucose concentration (Nathan et al., 2007;Nathan et al., 2008). Therefore, it is necessary to investigate the causal effects of these three T2D-related traits in the blood and how they differ in subsequent pathological disorders.

To efficiently identify causal associations between T2D-related traits and various phenotypes without potential biases or confounding and/or reverse causations, Mendelian randomization (MR) can be used to assess how genetic variants act as instruments for instrumental variable (IV) analysis aimed at estimating the causal effect of one trait on another. Using genetic variants as instruments, which are not associated with conventional confounders of observational studies, allows the MR approach to be considered analogous to randomized controlled trials (Burgess and Thompson, 2015). MR analysis requires three assumptions: 1) IVs are strongly associated with intermediate exposure, 2) IVs are independent of confounders, and 3) IVs affect outcomes only through the exposure path. If these assumptions hold, an inverse-variance-weighted (IVW) method provides the most efficient and unbiased estimates of causal effects (Burgess et al., 2020). Various MR methods have been proposed for providing a more robust approach under weaker assumptions (Burgess et al., 2013; Bowden et al., 2015; Bowden et al., 2016a; Bowden et al., 2016b; Verbanck et al., 2018).

The aim of this study was to assess the causal effect of T2D-related traits (FPG, 2h-PG, and HbA1c) on cancers and vascular diseases via MR analysis using several methods, including those measuring sensitivity in the MR-Base platform database (Hemani et al., 2016).

## 2 Materials and Methods

### 2.1 Exposure datasets

The exposure traits of interest were FPG, 2h-PG, and HbA1c. The summary statistics for T2D-related traits were obtained through large-scale genome-wide association study (GWAS) meta-analyses of 133,010 non-diabetic individuals from collaborating studies within the Meta-Analysis of Glucose and Insulin related traits Consortium (MAGIC) (Scott et al., 2012). In most of these studies, participants were of European ancestry and adults. A total of ~2.5 million genome-wide directly genotyped or imputed autosomal single-nucleotide polymorphisms (SNPs) were reported, including 36, 9, and 11 SNPs with genome-wide significant (*P* < 5 × 10^−8^) associations with FPG, 2h-PG, and HbA1c, explaining 4.8%, 1.7%, and 2.4% of the variance in the trait, respectively. Among these, SNPs were selected as IV candidates not in linkage disequilibrium (LD; *r*^2^<0.001) or within 10,000 kb of an established signal. To specify final IV sets, available genetic instruments for assessing outcome traits of interest were explored via the MR-Base platform database (https://www.mrbase.org/) or through the R package ‘TwoSampleMR’ (https://rdrr.io/github/MRCIEU/TwoSampleMR/). To reflect the same reference strand between exposure and outcome, alleles and effects were harmonized using effect/non-effect alleles and minor allele frequency for palindromic SNPs.

### 2.2 Outcome datasets

Human phenotypes were divided into two categories of diseases or traits known to be related to T2D. The first category was cancer at major sites: breast, gall bladder, lung [adenocarcinoma and squamous cell (SC) carcinoma], ovarian, pancreatic, and thyroid (differentiated types). The second category was vascular disease: coronary kidney disease (CKD), coronary artery disease (CAD), stroke, cardio-embolic stroke, small-vessel stroke, and high-density lipoprotein (HDL)/low-density lipoprotein (LDL) cholesterol levels. We obtained summary SNP-outcome associations with a total of 14 human health phenotypes through the MR-BASE platform. Additionally, information regarding each outcome trait of interest was extracted (e.g., author/study/consortium name, number of cases and controls, publication year, PubMed ID, study population, unit, etc.) and listed in Table 1.

**Table 1.** Description of data from MR-Base based on the phenotype

| Category | Trait | Consortium/<br>First author | PubMed ID | Unit | No. of<br>cases | No. of<br>controls | No. of<br>SNPs | Population |
| --- | --- | --- | --- | --- | --- | --- | --- | --- |
| 1. Cancer | Breast cancer | BCAC | 25751625 | LogOR | 15,748 | 18,084 | 13,011,123 | European |
|  | Lung cancer | ILCCO | 24880342 | LogOR | 11,348 | 15,861 | 8,945,893 | European |
|  | Lung cancer (SC) | ILCCO | 24880342 | LogOR | 3,275 | 150,038 | 8,893,750 | European |
|  | Ovarian cancer | OCAC | 28346442 | LogOR | 1,366 | 40,941 | 11,403,952 | European |
|  | Pancreatic cancer | PanScan1 | 19648918 | LogOR | 1,896 | 1,939 | 521,863 | European |
|  | Thyroid cancer | Kohler A | 23894154 | LogOR | 649 | 431 | 572,028 | European |
| 2. Vascular disease | CAD | VanderHarst P | 29212778 | LogOR | 122,733 | 424,528 | 7,934,254 | European |
|  | CKD | CKDGen | 26831199 | LogOR | 12,385 | 104,780 | 2,191,877 | Mixed |
|  | HDL cholesterol | GLGC | 24097068 | SD | — | 187,167 | 2,447,442 | Mixed |
|  | LDL cholesterol | GLGC | 24097068 | SD | — | 173,082 | 2,437,752 | Mixed |
|  | Stroke | Malik R | 29531354 | LogOR | 40,585 | 406,111 | 7,633,440 | European |
|  | (Cardio-embolic) | Malik R | 29531354 | LogOR | 7,193 | 406,111 | 8,271,294 | European |
|  | (Small-vessel) | Malik R | 29531354 | LogOR | 5,386 | 192,662 | 6,150,261 | European |

SC, squamous cell; SD, standard deviation; CKD, chronic kidney disease; CAD, coronary artery disease; LDL, low-density lipoprotein; HDL, high-density lipoprotein; logOR, log odds ratio.

### 2.3 MR assumptions

The assumptions of MR studies can be represented using causal directed acyclic graphs (DAG) (Figure 1). In a DAG, the genetic variant *G_j_* (*j*=1, 2, …,*J*) and the exposure, *X*, are denoted as *γ_j_*, and the association between the genetic variant, *G_j_*, and the outcome, *Y*, is denoted as *α_j_*. Associations between a confounding factor (*U*) and *G_j_*, *X*, and *Y* are denoted as *ψ_j_*, *K_x_*, and *K_y_*, respectively. In a two-sample MR setting, we refer to 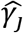 as an estimate from the *j*^th^ SNP-exposure association (with variance 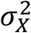) from sample 1 and 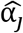 as an estimate from the *j*^th^ SNP-outcome association (with variance 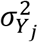) from sample 2.

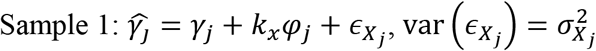

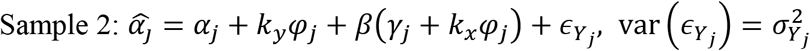

**Figure 1.**
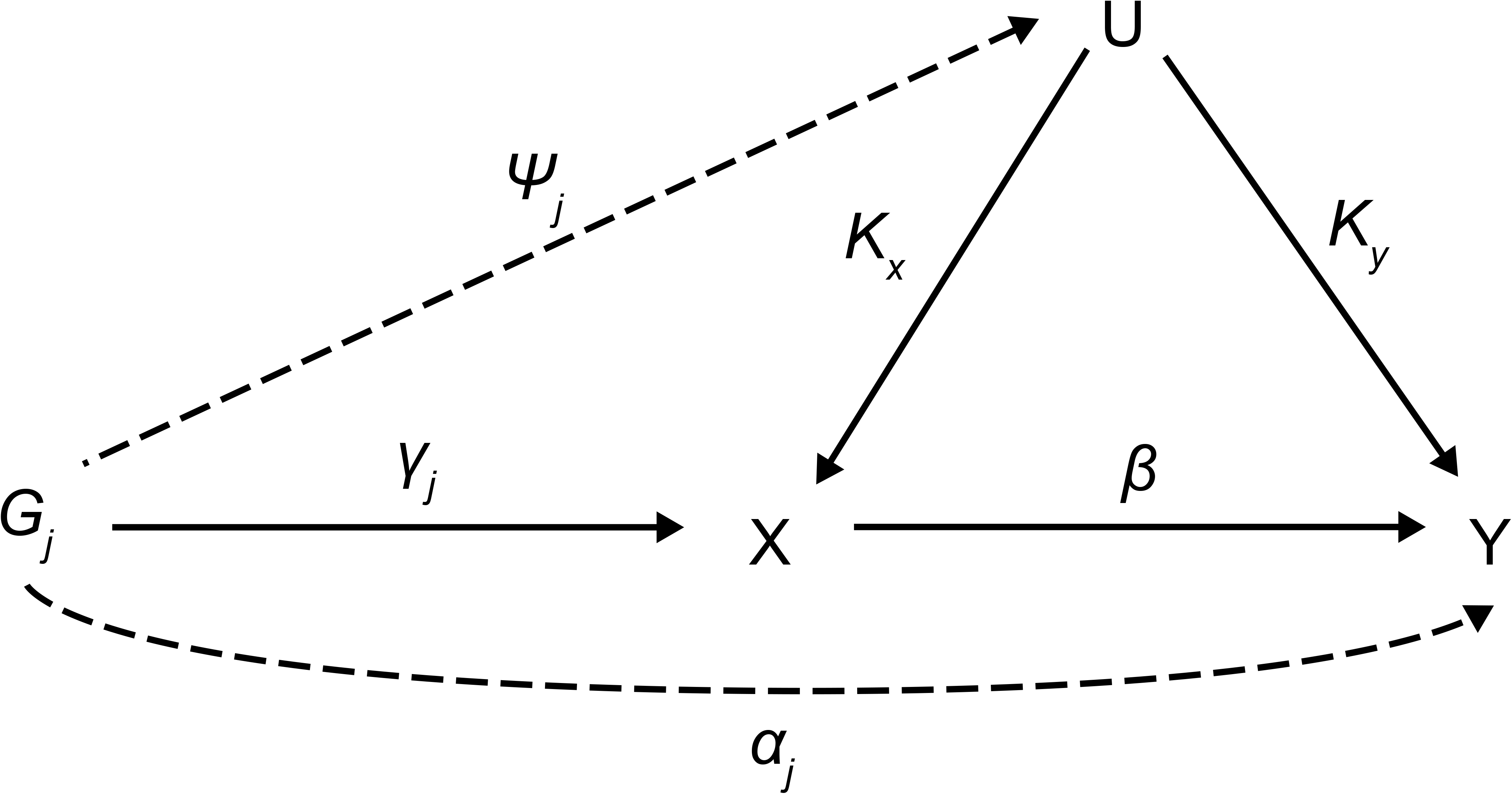
Causal directed acyclic graph for MR analysis. MR, Mendalian randomization.

The genetic variant, *G_j_*, for valid IVs must satisfy the following three core assumptions; (i) IV1: *γ_j_ ≠* 0, (ii) IV2: *φ_j_* = 0, (iii) IV3: *α_j_* = 0. Furthermore, MR requires a “NO Measurement Error” (NOME) assumption and an Instrument Strength Independent of Direct Effect (InSIDE) assumption. It is important to assess the instrument strength to prevent weak instrument bias on MR analysis. We evaluated weak instruments with mean F-statistics, and the degree of violation of the NOME assumption was quantified using the previously reported *I^2^* statistic (ranging 0–1) (Bowden et al., 2016b). Higher values for *I^2^* indicate lesser dilution of the causal effect estimate.

### 2.4 MR methods

Using all genetic variants, *G_j_*, that satisfy the three IV assumptions and the NOME and InSIDE assumptions, the causal effect of exposure on the outcome can be consistently estimated from the ratio estimates and averaged using an IVW method (Burgess et al., 2013). The IVW estimate is the most efficient method when all genetic variants satisfy all three IV assumptions. Cochran’s Q statistic was used to quantify heterogeneity (Greco et al., 2015; Bowden et al., 2017).

However, the estimate could be biased if one or more variants are invalid. The weighted median method provides valid causal estimates, even if up to 50% of the instruments are invalid. The median is unaffected by outliers, making the weighted median estimate insensitive to a pleiotropic genetic variant. Causal effects are obtained from the weighted median of the ratio estimates in genetic instruments, resulting in smaller standard errors receiving more weight (Bowden et al., 2016a).

The MR-Egger method allows all SNPs to be used as invalid instruments but requires variants to satisfy the InSIDE assumption, enabling estimation of appropriate causal effects in the presence of pleiotropic effects (Bowden et al., 2015). This model is suitable for linear regression and the intercept term, *β_0E_*, is interpreted as the average horizontal pleiotropic effect across the genetic variants (Bowden et al., 2015). Rucker’s Q’ statistic from MR-Egger was used to quantify directional heterogeneity (Greco et al., 2015; Bowden et al., 2017). If estimates of *β_0E_* equal zero, the MR-Egger slope estimate will be the same as the IVW estimate (Burgess and Thompson, 2015). However, when the *I^2^* statistic quantifying the strength of NOME violation for IVs for the MR-Egger method is low, a magnitude of regression dilution still occurs. In cases where the NOME assumption is violated, the SIMEX method can be used to correct attenuation bias (Bowden et al., 2016b).

Violation of IV3 (no horizontal pleiotropy) can raise a severe bias in MR analysis. The MR-PRESSO test has an advantage over MR-Egger, in that it identifies and removes pleiotropic SNPs. The test comprises three parts: 1) the MR-PRESSO global test detects horizontal pleiotropy, 2) the outlier-corrected causal estimate corrects for the detected horizontal pleiotropy, and 3) the MR-PRESSO distortion test estimates whether the causal estimates differ significantly (*P* < 0.05) following adjustment for the outliers (Verbanck et al., 2018). Therefore, MR-PRESSO results are preferable in the presence of a horizontal pleiotropic effect.

The appropriate methods differ according to the assumptions satisfied, and the most suitable choices are presented in Table 2 and 3. The IVW method is the most efficient way to estimate the causal effect when all genetic variants are valid instruments (Burgess et al., 2020). In cases where the MR assumption of no pleiotropy is not met, the MR-PRESSO test detects possible outliers and provides consistent estimates following outlier removal (Burgess and Thompson, 2017). When some genetic variants are invalid (<50%), the weighted median approach can be used as an alternative method of providing a consistent estimate (Bowden et al., 2016a). By contrast, MR-Egger can obtain a causal estimate by correcting directional pleiotropy but has the disadvantage of low power (Bowden et al., 2015). If the NOME assumption is violated (*I*^2^ < 90%), the MR-Egger (SIMEX) method would be suitable (Bowden et al., 2016b).

**Table 2.** Recommended MR methods by assumption of IVs

| No weak IVs (F>10) | NOME | No Heterogeneity | Recommended Methods |
| --- | --- | --- | --- |
| Satisfied | Satisfied | Satisfied | IVW |
| Satisfied | Satisfied | At least one violated | →Table 3 |
| Satisfied | Violated | Satisfied | IVW |
| Satisfied | Violated | At least one violated | MR-Egger (SIMEX) |
| Violated | Satisfied | Satisfied | Weighted median |
| Violated | Satisfied | At least one violated | →Table 3 |
| Violated | Violated | Satisfied | MR-Egger (SIMEX) |
| Violated | Violated | At least one violated | MR-Egger (SIMEX) |

IV, instrument variable; NOME, NO Measurement Error; IVW, inverse–variance–weighted; SIMEX, simulation extrapolation;

**Table 3.** Recommended MR methods by heterogeneity test

| Q test<sup>†</sup> | Q' test<sup>†</sup> | MR-PRESSO global test<sup>‡</sup> | Recommended Methods |
| --- | --- | --- | --- |
|  |  |  | IVW |
| ✓ | ✓ |  | MR-Egger |
|  | ✓ |  | IVW |
| ✓ |  |  | MR-Egger |
|  |  | ✓ | IVW, if there is no weak IV<br>MR-PRESSO, otherwise |
| ✓ | ✓ | ✓ | MR-PRESSO |
|  | ✓ | ✓ | IVW, if there is no weak IV (F>10)<br>MR-PRESSO, ow (F<10) |
| ✓ |  | ✓ | MR-Egger, MR-PRESSO |

✓: significant (heterogeneity exists)
<sup>†</sup>Greco et al., 2015; Bowden et al., 2017
<sup>‡</sup> Verbanck et al., 2018
IV, instrument variable; IVW, inverse–variance–weighted; MR-PRESSO, MR-Pleiotropy RESidual Sum and Outlier

### 2.5 Bidirectional MR analysis

We conducted bidirectional MR analysis to investigate the presence of reverse causality among associations between T2D-related traits and outcomes of interest. This was performed by switching the exposure and outcomes in opposite directions.

### 2.6 MR power analysis

Power calculations were conducted at https://sb452.shinyapps.io/power/ (Burgess et al., 2020). The proportion of variance in the exposure explained by the genetic variants (*R^2^*) were required for MR power analysis, with 0.048 (FPG), 0.017 (2h-PG), and 0.024 (HbA1c) used, respectively. We assumed odds ratios (ORs) of 1.1 and 1.2 for binary outcomes and changes in outcomes in standard deviation (SD) units per SD change in exposure (0.1 and 0.2) for continuous outcomes. Statistical power evaluations at the conservative significance level [0.007 (Bonferroni correction with 7 tests)] are plotted in Figure 2.

**Figure 2.**
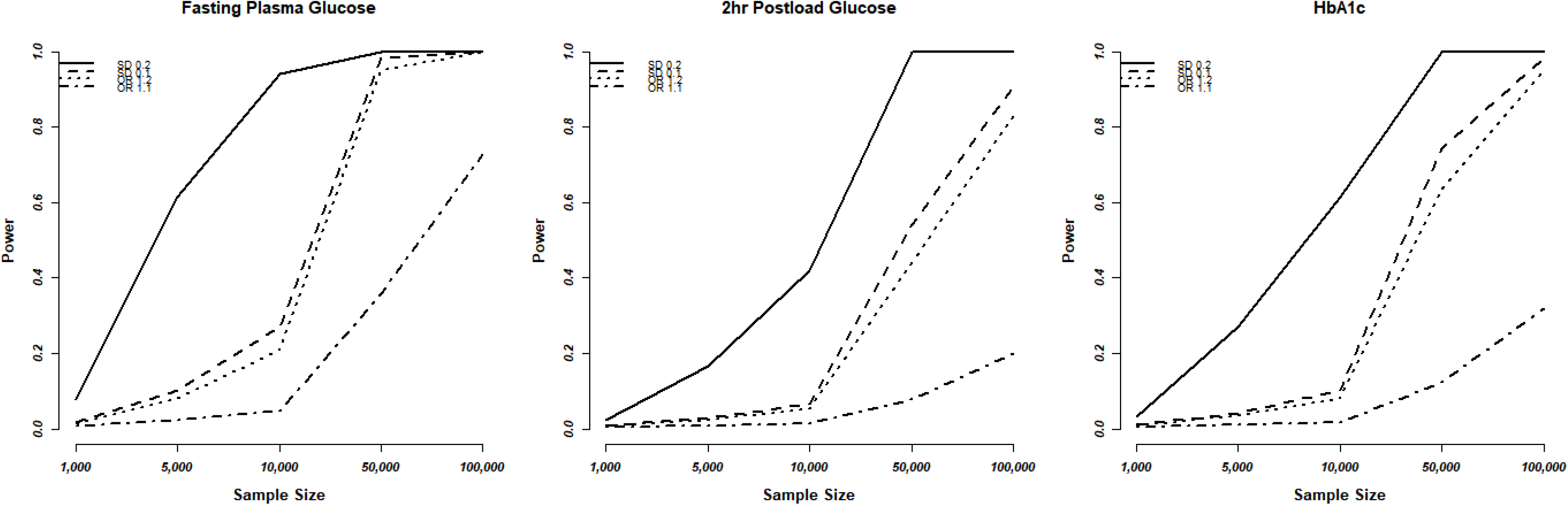
Statistical power evaluations of MR analyses based on the T2D-diagnosis criteria. (A) FPG, (B) 2h-PG, and (C) HbA1c. We used a conservative significance threshold of *P* < 0.007 with Bonferroni correction using 7 of testing. MR, Mendalian randomization; T2D, type 2 diabetes; FPG, fasting plasma glucose; 2h-PG, 2-h plasma glucose; HbA1c, hemoglobin A1c; OR, odds ratio; SD, standard deviation.

## 3 Results

A total of 34, 7, and 11 genetic variants associated with FPG, 2h-PG, and HbA1c respectively, were available as potential instruments from studies included in MAGIC. Each IV set showed genome-wide significant (*P* < 5 × 10^−8^) associations with T2D-related traits and were not in LD or within 10,000 kb of an established signal. To investigate IV quality, we generated F-statistics, *I*^2^ values, and *P*-values for Cochran’s Q statistic from IVW, Rucker’s Q’ statistic from MR-Egger, and MR-PRESSO global test (Table 4). All instruments used for MR analyses had F-statistics >10, indicating no evidence of weak instrument bias. Rejection of the null hypothesis of the Cochran’s Q statistic for heterogeneity suggested potential pleiotropy in the genetic variants and did not indicate that the InSIDE assumptions were invalid. When the pleiotropic effect was present, MR-Egger (with and without SIMEX), and MR-PRESSO) were performed rather than using the IVW method. The instruments corresponding to FPG satisfied the NOME assumption (*I*^2^ > 90) but only partially satisfied this for HbA1c (*I^2^* > 90 in only some cases) and did not satisfy this in the case of 2h-PG (*I*^2^< 90). When the NOME assumption was violated, the results of MR-Egger (SIMEX) were generated. Using these IVs, we performed MR analyses for a total of 14 human health phenotypes, with all results (3 exposures × 13 phenotypes × 5 methods = 185 results) presented in Supplementary Table 1. Application of Bonferroni correction to each disease category (0.05/6 = 0.008 for cancer; and 0.05/7 = 0.007 for vascular disease) revealed two significant phenotypes (CAD and LDL level) associated with T2D-related traits (Table 5). Additionally, we confirmed these relationships through bidirectional and replication analyses (Tables 5 and 6).

**Table 4.** Assumption check for instrumental variables

| | FPG ($R^2 = 0.048$) | | | | 2h-PG ($R^2 = 0.017$) | | | | HbA1c ($R^2 = 0.024$) | | | |
| --- | --- | --- | --- | --- | --- | --- | --- | --- | --- | --- | --- | --- |
| | N | F | $I^2$<br>(%) | Q<br>Q'<br>RSS | N | F | $I^2$<br>(%) | Q<br>Q'<br>RSS | N | F | $I^2$<br>(%) | Q<br>Q'<br>RSS |
| **1. Cancer** |  |  |  |  |  |  |  |  |  |  |  |  |
| Breast cancer | 34 | 130.5 | 96.8 | <0.001<br><0.001<br><0.001 | 7 | 43.5 | 29.8 | <0.001<br>0.019<br>0.023 | 11 | 77.6 | 92.2 | 0.110<br>0.249<br>0.371 |
| Lung cancer | 33 | 133.0 | 97.3 | 0.182<br>0.183<br>0.211 | 7 | 43.5 | 36.2 | 0.322<br>0.397<br>0.258 | 11 | 77.6 | 87.6 | 0.021<br>0.063<br>0.089 |
| SC lung cancer | 33 | 43.5 | 40.1 | 0.599<br>0.553<br>0.477 | 7 | 43.5 | 40.1 | 0.875<br>0.786<br>0.913 | 11 | 77.6 | 87.9 | 0.198<br>0.154<br>0.216 |
| Ovarian cancer | 33 | 133.5 | 97.4 | 0.530<br>0.490<br>0.412 | 7 | 43.5 | 28.1 | 0.323<br>0.224<br>0.362 | 11 | 77.6 | 87.8 | 0.243<br>0.334<br>0.298 |
| Pancreatic cancer | 25 | 111.7 | 96.4 | 0.138<br>0.122<br>0.199 | 6 | 45.0 | 31.8 | 0.710<br>0.594<br>0.642 | 8 | 82.1 | 90.9 | 0.239<br>0.186<br>0.214 |
| Thyroid cancer | 27 | 144.9 | 97.9 | 0.183<br>0.248<br>0.275 | 5 | 47.4 | 21.9 | 0.434<br>0.703<br>0.621 | 9 | 81.7 | 91.2 | 0.882<br>0.899<br>0.885 |
| **2. Cardiovascular disease** |  |  |  |  |  |  |  |  |  |  |  |  |
| CAD | 34 | 130.5 | 97.3 | <0.001<br><0.001<br><0.001 | 7 | 43.5 | 23.0 | <0.001<br><0.001<br><0.001 | 11 | 77.6 | 88.0 | 0.002<br>0.014<br>0.018 |
| CKD | 32 | 135.8 | 97.4 | 0.106<br>0.114<br>0.151 | 7 | 43.5 | 29.1 | 0.482<br>0.451<br>0.395 | 10 | 82.1 | 86.5 | 0.632<br>0.548<br>0.421 |
| HDL cholesterol | 34 | 130.5 | 97.3 | <0.001<br><0.001<br><0.001 | 7 | 43.5 | 2.4 | <0.001<br><0.001<br><0.001 | 11 | 77.6 | 90.9 | 0.545<br>0.589<br>0.436 |
| LDL cholesterol | 34 | 130.8 | 97.2 | <0.001<br><0.001<br><0.001 | 7 | 43.5 | 5.7 | <0.001<br><0.001<br><0.001 | 11 | 77.6 | 91.1 | <0.001<br><0.001<br><0.001 |
| Stroke | 34 | 130.5 | 97.3 | <0.001<br><0.001<br><0.001 | 7 | 43.5 | 39.9 | 0.069<br>0.051<br>0.067 | 11 | 77.6 | 86.0 | 0.075<br>0.084<br>0.092 |
| (Cardio-embolic) | 34 | 130.5 | 97.4 | 0.498<br>0.649<br>0.557 | 7 | 43.4 | 35.2 | 0.733<br>0.611<br>0.694 | 11 | 77.6 | 88.7 | 0.255<br>0.247<br>0.275 |
| (Small-vessel) | 32 | 135.8 | 97.6 | 0.038<br>0.130<br>0.199 | 7 | 43.5 | 55.6 | 0.675<br>0.636<br>0.645 | 10 | 76.88 | 86.6 | 0.556<br>0.522<br>0.512 |

F, F-statistic; Q, $P$ -value for the Q-test from IVW; Q', $P$ -value for the Q'-test from MR-Egger; RSS, $P$ -value for MR-PRESSO global test, SC, squamous cell; CKD, chronic kidney disease; CAD, coronary artery disease; LDL, low-density lipoprotein; HDL, high-density lipoprotein; FPG, fasting plasma glucose; 2h-PG, 2-h plasma glucose; HbA1c, hemoglobin A1c.

**Table 5.**
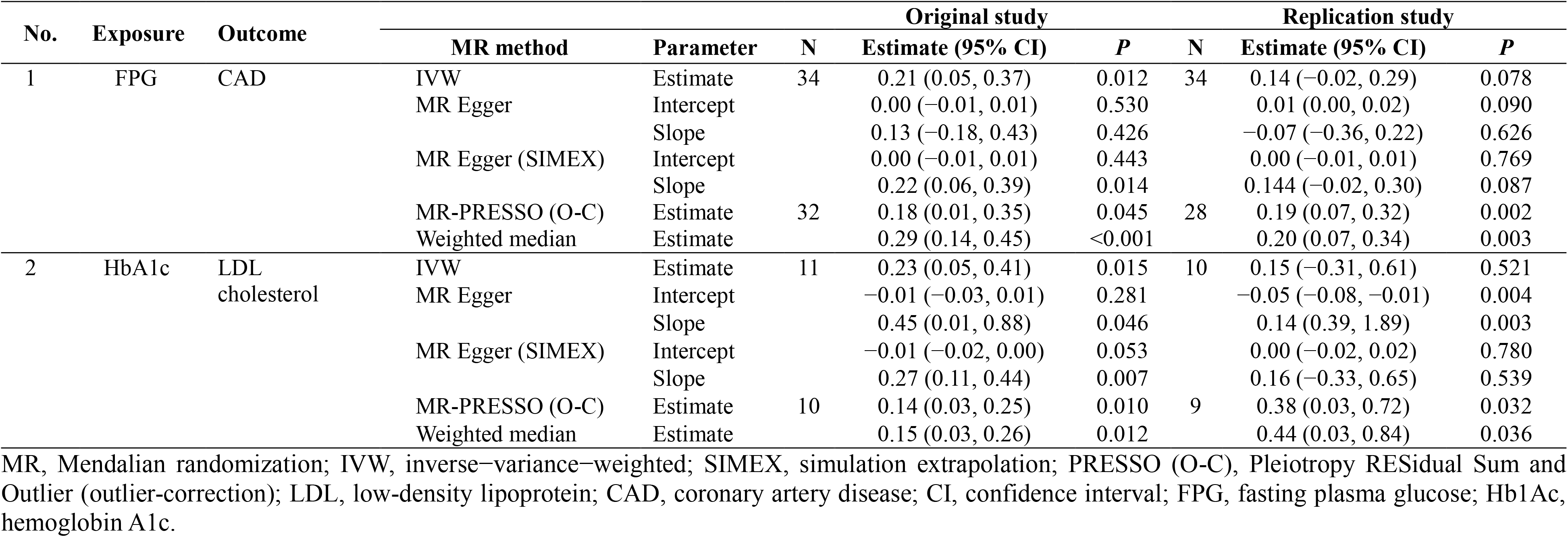
Significant results from MR and replication analyses

**Table 6.**
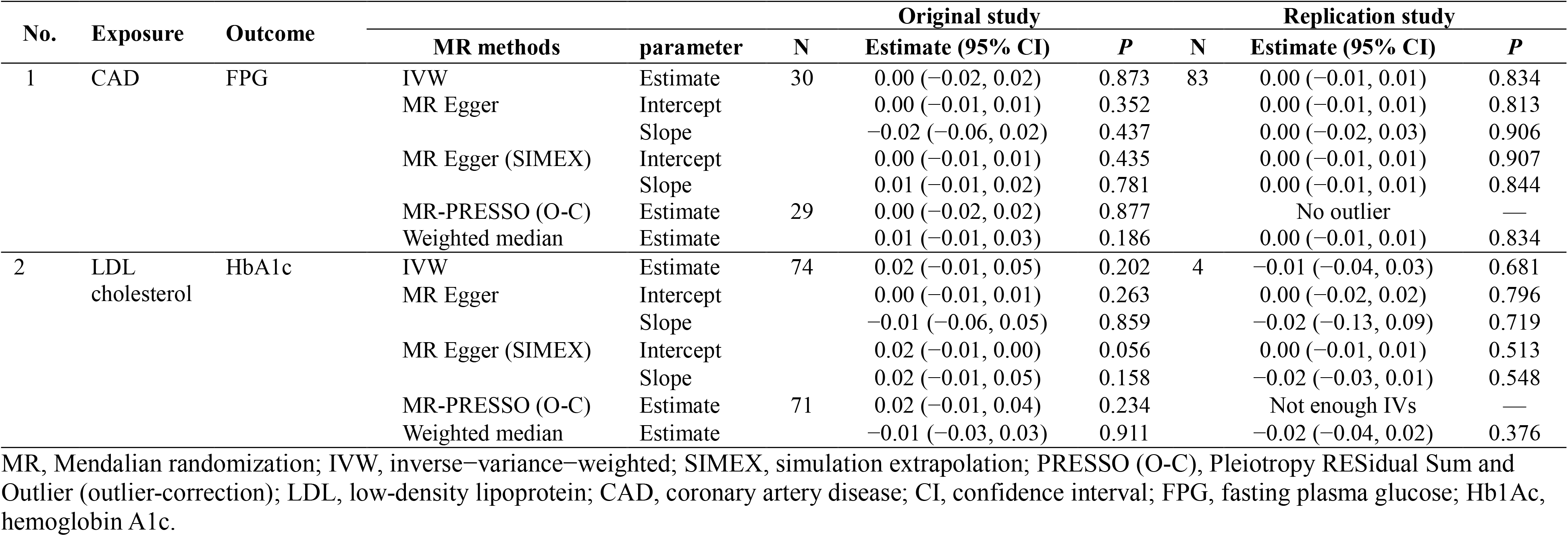
Significant results from bidirectional MR analysis

### 3.1 T2D-related traits and cancers

We considered FPG, 2h-PG, and HbA1c as exposure traits. For FPG, IVs for lung, ovarian, pancreatic, and thyroid cancer satisfied the IV assumptions (F statistics >10, *I^2^* > 90, Q-P>0.05), and IVW was selected for MR analyses (Table 3). No significant causal association was observed between FPG and lung (*P*=0.721), ovarian (*P*=0.632), pancreatic (*P*=0.768), and thyroid (*P*=0.612) cancer. A pleiotropic effect was observed in breast cancer through Q (*P*<0.001), Q’ (*P*<0.001) statistics, and the MR-PRESSO global test (*P*<0.001), and MR-PRESSO did not yield significant outcomes (*P*=0.364). The NOME assumption was violated in SC lung cancer (*I*^2^ <90), and the MR-Egger (SIMEX) method was used. The MR-Egger (SIMEX) method yielded nominally significant (*P*<0.05) causal effects (*P*=0.032). Furthermore, when 2h-PG was considered an exposure trait, IVs for all cancers, except for breast cancer, we found no weak instrument bias (F>10) and no heterogeneity (Q-P>0.05, Q’-P>0.05, MR-PRESSO global test-*P*>0.05), and the IVW method was used. However, IVs for breast cancer have a measurement error (*I^2^* <90), and the MR-Egger (SIMEX) method was used. None of the IVs were significant for breast (*P*=0.303), lung (*P*=0.721), SC lung (*P*=0.037), ovarian (*P*=0.632), pancreatic (*P*=0.768), and thyroid (*P*=0.612) cancer. Moreover, regarding HbA1c, evidence of violations of IV assumptions for all cancers was obtained (F statistics >10, Q-P>0.05, Q’-*P*>0.05, MR-PRESSO global test-*P*>0.05), and IVW was applied. No significant association was observed between HbA1c and breast (*P*=0.922), lung (*P*=0.173), SC lung (*P*=0.115), ovarian (*P*=0.719), pancreatic (*P*=0.374), and thyroid (*P*=0.417) cancer.

For lung, breast, and ovarian cancer, we assumed an OR of 1.2 and we determined the statistical power at between 40% and 70%. The highest power was observed for FPG with the highest *R*^2^, followed by HbA1c and 2h-PG. The estimated statistical power was the highest (>80%) for SC lung cancer for all T2D-related traits owing to a sample size of >100,000 individuals if the standardized effect size is assumed to be same. However, for pancreatic and thyroid cancers, the sample size was small (3,835 and 1,080, respectively), thus decreasing the statistical power, indicating the possibility of false-negative results. The overall estimated power (Figure 2) revealed no causal effect of FPG, 2h-PG, and HbA1c on breast, lung, SC lung, ovarian, pancreatic, and thyroid cancers (*P* < 0.008 after Bonferroni correction) (Supplementary Table 1).

### 3.2 T2D-related traits and vascular diseases

All data for vascular diseases were from a sample size of >100,000 patients, giving them a power of ≥80%, except for detecting an OR of 1.1. We found no causal effect of FPG, 2h-PG, or HbA1c on CKD, HDL level, stroke, or stroke subtype, but two significant causal relationships were observed for FPG with CAD and HbA1c with HDL level. Interestingly, three T2D-related traits used as criteria for diagnosing T2D showed different results for the same phenotype. First, on using FPG as an exposure trait, IVs for CKD and cardio-embolic stroke strongly satisfied the IV assumptions (F statistics >10, *I^2^* > 90, F statistics >10, Q-P>0.05, Q’-P>0.05, MR-PRESSO global test-P>0.05), and the IVW approach was selected (Table 2). However, FPG had no causal effects on CKD (*P*=0.351) and cardio-embolic stroke (*P*=0.118) were observed, but was nominally significant on small-vessel stroke (*P*=0.025). In the case of CAD, HDL/LDL cholesterol, and stroke, we found heterogeneity (Q-*P*<0.05, Q’-*P*<0.05, MR-PRESSO global test-*P*<0.05), and the MR-PRESSO method was applied. Nominally significant results were observed for CAD (MR-PRESSO *P*=0.045) and non-significant results were observed for HDL cholesterol (*P*=0.265), LDL cholesterol (*P*=0.225), and stroke (*P*=0.135). Second, when 2h-PG was used as an exposure trait, IVs for CKD, stroke, cardio-embolic stroke, and small-vessel stroke strongly satisfied the IV assumptions (F statistics >10, F statistics >10, Q-*P*>0.05, Q’-*P*>0.05, MR-PRESSO global test-*P*>0.05), and the IVW method used. Non-significant causal effects were observed for 2h-PG on CKD (*P*=0.183), stroke (*P*=0.338), cardio-embolic stroke (*P*=0.530), and small-vessel stroke (*P*=0.084). In the case of CAD, HDL/LDL cholesterol, all have measurement error (*I*^2^ <90) with heterogeneity ((Q-*P*<0.05, Q’-*P*<0.05, MR-PRESSO global test-*P*<0.05), and the MR-Egger (SIMEX) method was used. Non-significant causal effects were observed for 2h-PG on CAD (*P*=0.301), HDL cholesterol (*P*=0.074), and LDL cholesterol (*P*=0.241). Third, when HbA1c was considered an exposure trait, IVs for CKD, HDL cholesterol, stroke, cardio-embolic stroke and small-vessel stroke strongly satisfied the IV assumptions (F statistics >10, Q-*P*>0.05, Q’-*P*>0.05, MR-PRESSO global test-*P*>0.05), and the IVW method was selected. However, no causal effects of HbA1c were observed on CKD (*P*=0.337), HDL cholesterol (*P*=0.206), and stroke (*P*=0.567), but there were nominally significant implications for cardio-embolic stroke (*P*=0.023) and small-vessel stroke (*P*=0.046). Owing to the heterogeneity in CAD and LDL cholesterol (Q-*P*<0.05, Q’-*P*<0.05, MR-PRESSO global test-*P*<0.05), the MR-PRESSO method was considered, and nominally significant results were obtained for LDL cholesterol (*P*=0.010), but non-significant for CAD (*P*=0.069).

Significant effects were found for FPG-CAD and HbA1c-LDL cholesterol. Regarding FPG-CAD, all SNP-exposure and SNP-outcome effects are presented in Supplementary Table 2. We found two SNPs significantly correlated with CAD (rs1260326: *P* = 2.40 × 10^−5^; and rs7651090: *P* = 1.20 × 10^−5^); however, given that they exhibited balanced pleiotropy, they were not excluded from the analysis (but were excluded from MR-PRESSO tests). A generated funnel plot showed symmetry, indicating heterogeneity due to horizontal pleiotropy (Figure 3A). The associations of the variants with FPG and CAD are shown in a scatter plot with five MR-fitted lines (Figure 3B). Thirty-four SNPs were considered instruments, and no weak instrument bias was noted with no violation of NOME assumption, albeit with heterogeneity. Therefore, we assessed the other several sensitivity methods, and observed causal effects of CAD on FPG from the weighted median (*P*<0.001). Moreover, we verified that reverse causality did not exist (Table 6). In the replication study using the same IVs and different GWAS data for outcome (PmID = 29212778, N = 296,525, P = European, and unit = logOR), there was no weak instrument bias of IVs (N=34, F statistics 43.5) but the heterogeneity assumption was violated (Q-*P*<0.05, Q’-*P*<0.05, MR-PRESSO global test-*P*<0.05). Therefore, MR-PRESSO was selected, and we found that FPG has a positively causal effect on CAD in accordance with the MR-PRESSO method (*P* = 0.002) (Table 5). On bidirectional MR analysis in the replication study, 83 SNPs were considered instrument variables. Weak instrument bias (F-statistics 77.2) and the NOME assumption (*I^2^* =92.4) were preserved; however, heterogeneity was observed Q-*P*<0.05, Q’-*P*<0.05, MR-PRESSO global test-*P*<0.05). MR-PRESSO revealed no causal effect of CAD on FPG (Table 6).

**Figure 3.**
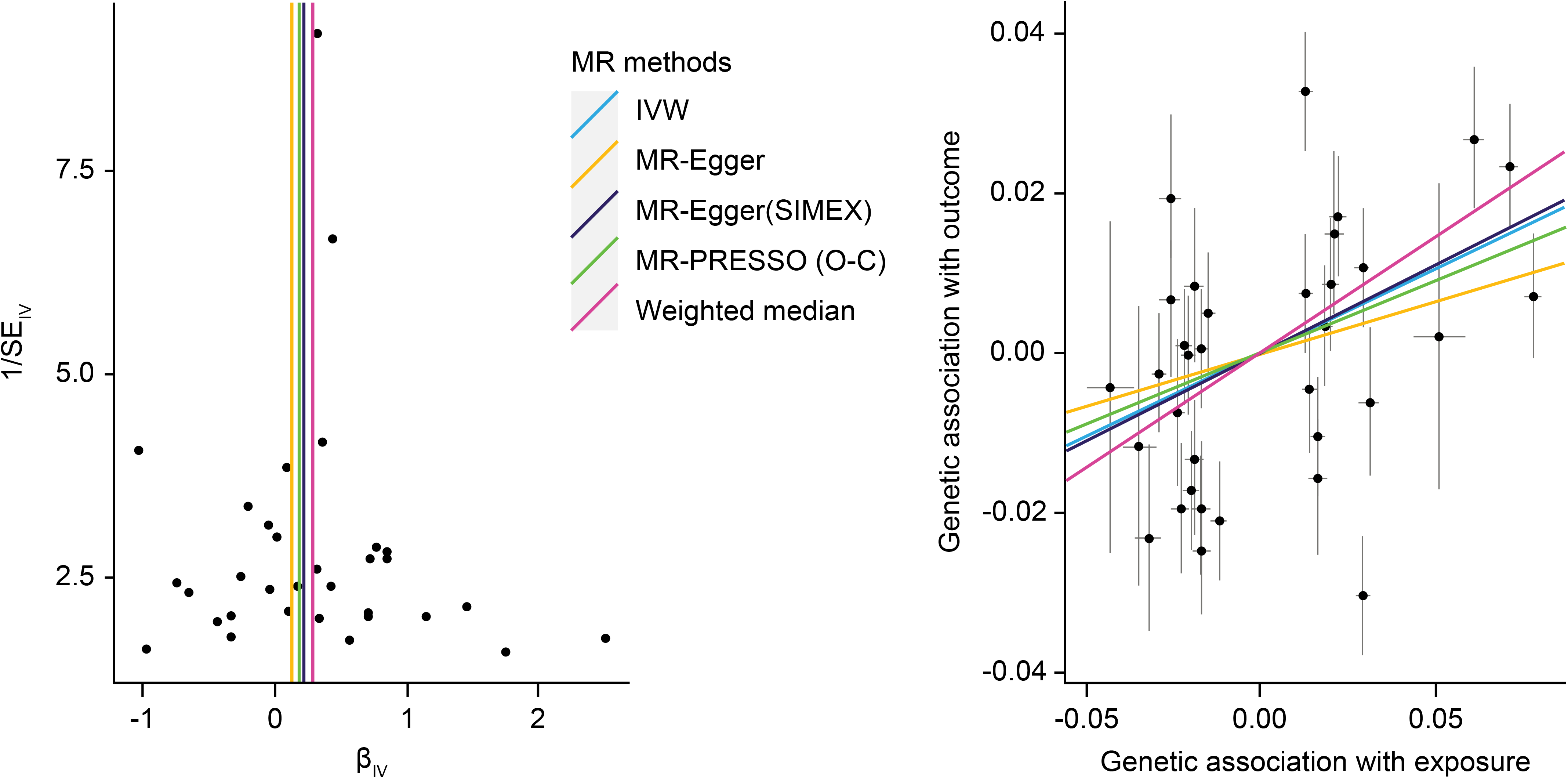
MR analysis of the effect of FPG on CAD. (A) Funnel plot displaying individual causal effect estimates for FPG on CAD. Dots representing the estimated causal effect for each IV. (B) The association between the effect size estimates on the FPG (X-axis) and CAD (Y-axis) for all SNPs that served as IVs. FPG, fasting plasma glucose; CAD, coronary artery disease; IV, instrumental variable; SNP, single-nucleotide polymorphism; SIMEX, simulation extrapolation; PRESSO (O-C), Pleiotropy RESidual Sum and Outlier (outlier-correction).

Regarding HbA1c and LDL cholesterol, SNP-exposure and SNP-outcome effects (Supplementary Table 3) indicated that one SNP was significantly correlated with LDL level (rs1800562; *P*=4.42 × 10^−4^), with this SNP excluded from MR-PRESSO analysis. Figure 4A shows a funnel plot indicating slight non-symmetry, suggesting the presence of heterogeneity due to horizontal pleiotropy. The scatter plot in Figure 4B shows the associations of the variants with HbA1c and LDL level. Eleven SNPs were considered instruments, and no weak instrument bias was noted with no violation of NOME assumption, albeit with heterogeneity. Therefore, we assessed several other sensitivity methods, and observed causal effects of HbA1c on LDL cholesterol from MR-Egger (SIMEX) (*P*=0.007). In addition, reverse causality was not identified (Table 6). Seventy-four SNPs were considered instruments, and no weak instrument bias was noted (F-statistics 153.9), with no violation of the NOME assumption (*I^2^* =97.7). However, heterogeneity was observed (Q-*P*<0.05, Q’-*P*<0.05, MR-PRESSO global test-*P*<0.05), and MR-PRESSO revealed no causal effect of LDL cholesterol on HbA1c (*P*=0.234). Replication analysis using the same IVs and different GWAS data for the outcome-SNP effect (pmID = 28887542, N = 9,961, P = European, unit = mg/dL) revealed no evidence of a weak instrument bias (N=11, F-statistics 77.6) and no heterogeneity (Q-*P*>0.05, Q’-*P*>0.05, MR-PRESSO global test>0.05), but the NOME assumption (*I^2^* =87.9) was violated. Therefore, MR-PRESSO revealed significant results for the causal effect of HbA1c on LDL cholesterol (*P*=0.032) (Table 5). On bidirectional MR analysis for the replication study, 4 SNPs were considered instrument variables. No weak instrument bias (F-statistics 42.9) and no heterogeneity (Q-*P*>0.05, Q’-*P*>0.05, MR-PRESSO global test>0.05) were observed; however, a violation of the NOME assumption (*I^2^* =4) was noted. Accordingly, IVW was considered, and no causal effect of LDL cholesterol on HbA1c was observed (*P*=0.681) (Table 6).

**Figure 4.**
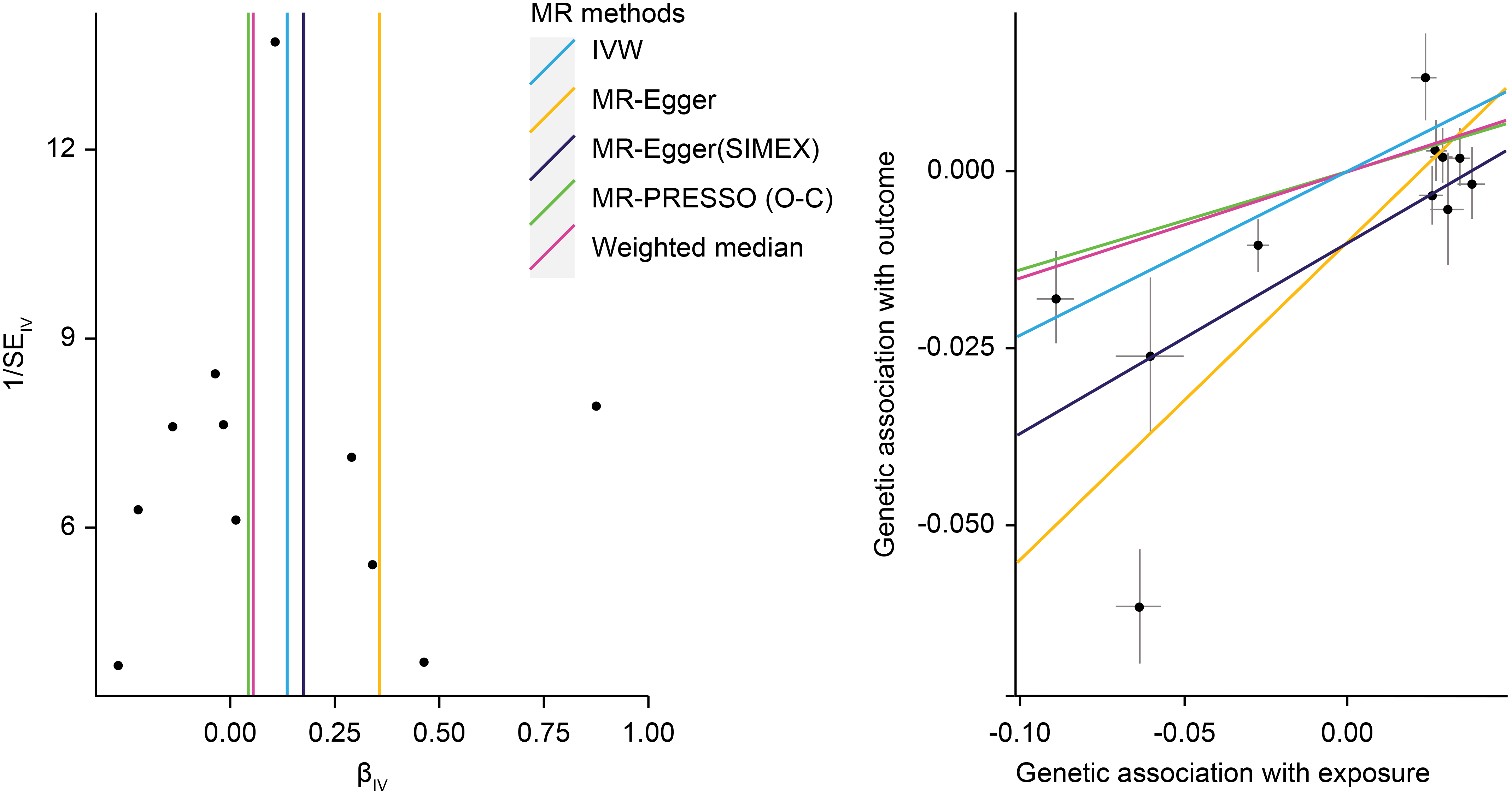
MR analysis of the effect of HbA1c on LDL levels. (**A**) Funnel plot displaying individual causal effect estimates for HbA1c on LDL levels. Dots represent the estimated causal effect for each IV. (**B**) The relationship between the effect size estimates on HbA1c (X-axis) and LDL level (Y-axis) for all SNPs that served as IVs. MR, Mendalian randomization; HbA1c, hemoglobin A1c; LDL, low-density lipoprotein; IV, instrumental variable; SNP, single-nucleotide polymorphism; SIMEX, simulation extrapolation; PRESSO (O-C), Pleiotropy RESidual Sum and Outlier (outlier-correction).

## 4 Discussion

In this study, we performed MR analysis of the effect of T2D-related traits on 13 human health phenotypes using GWAS results and data from the MR-BASE registry. In particular, MR analysis was conducted according to three T2D-related criteria (FPG and 2h-PG from the OGTT and HbA1c). MR analyses reduce potential confounding effects and reverse causation, and our results are concurrent with those of previous epidemiological studies. Previous large meta-analyses or systematic reviews of epidemiological studies show that the association between T2D and cancer development is unclear (Tsilidis et al., 2015). Moreover, most epidemiological studies report limitations in findings of T2D-related association with cancers, because they were based on self-reported health assessments with high specificity (>90%) but low sensitivity (66%) as compared with medical records (Okura et al., 2004). Recently, results of MR analysis indicated no strong evidence supporting a causal relationship between T2D and major solid tumors (stomach, colorectal, liver, pancreas, lung, breast, and prostate) (Goto et al., 2020). Similarly, in the present study, analysis of European data from the MR-Base registry revealed no significant causal effect of T2D-related traits on breast, lung, SC lung, ovarian, pancreatic, and thyroid cancers. Although T2D and cancer share a number of risk factors, such as hyperglycemia, insulin resistance, and dyslipidemia, a relationship between the diseases has not been fully demonstrated (Vigneri et al., 2009). Additionally, studies have reported correlations between hypoglycemic agents and cancer incidence, although these findings remain controversial (Alimova et al., 2009;Currie et al., 2009).

In T2D patients, the risk of death from cardiovascular disease increases along with elevated FPG and HbA1c levels, with HbA1c level correlated with microvascular and microvascular complications (Kannel and McGee, 1979;Group, 1998;Okura et al., 2004). Therefore, hyperglycemia represents a strong independent factor for cardiovascular disease, with the risk increasing 2- to 3-fold in men and 3- to 4-fold women diagnosed with T2D relative to those without T2D (Kannel and McGee, 1979;Okura et al., 2004). A longitudinal study involving follow-up for 8 years of 2,363 non-diabetic adults between the ages of 50 and 75 years reported significant association between 2h-PG and HbA1c levels and an increased risk of death from cardiovascular disease (De Vegt et al., 1999). Moreover, that study identified HbA1c level as not only predictive of improved better mortality from cardiovascular disease relative to FPG and 2h-PG (Park et al., 1996) but also an independent risk factor for atherosclerosis and cardiovascular disease independent of T2D (Nakamura et al., 1993;Kanauchi et al., 2001). In the present study, our findings indicated that vascular disease and LDL level were significantly linked with HbA1c level but not FPG or 2h-PG.

We found that different characteristics related to FPG, 2h-PG, and HbA1c differentially influenced IV characteristics. The 2h-PG results from an OGTT represent a standard test for T2D diagnosis. Although 2h-PG testing is more highly sensitive and specific than FPG testing, its low reproducibility is a disadvantage (Peters et al., 1996). The low reproducibility is a consequence of changes in 2-h glucose concentrations for each measurement within a 48-h or 1-week time period in the same individual. On the other hand, FPG testing is simple and reproducible; however, the sensitivity for T2D diagnosis is poor, because it does not allow accurate identification of hyperglycemia after glucose load (Davidson et al., 1999). HbA1c reflects overall tissue protein glycation and can better reflect the overall biological effect of blood sugar as a 3-month average blood sugar estimate (Peterson et al., 1998); however, HbA1c measurements can be affected by hemoglobin disease, chronic renal failure, testing methods, and/or specific dosage (Barr et al., 2002). Therefore, these findings suggest that the measurement error associated with SNP-exposure associations might be large when using any of these criteria. A previous study showed that calculation of the *I*^2^ value confirmed the inadequacy of the NOME assumption due to measurement error related to 2h-PG testing (Bowden et al., 2016b). Furthermore, reports indicated that the HbA1c level shows less variability in day-to-day within-person variance than FPG (<2% for HbA1c vs. 12–15% for FPG) (Ollerton et al., 1999), and the intra-individual coefficient of variation for FPG (6.4%) is less than that for 2h-PG (16.7%) (Mooy et al., 1996). Therefore, MR analysis using 2h-PG as an exposure can be expected to increase the reliability of MR-Egger (SIMEX) findings relative to other methods. In the cases of FPG and HbA1c, IVW results and the sensitivity analysis methods should be examined more broadly.

We performed MR analysis using public data from previous large-scale GWAS studies. Producing in-house genetic data is expensive and requires substantial human resources, making it difficult for many individual researchers lacking access to appropriate datasets. A two-sample MR approach represents an effective method for discovering novel causal relationships through the use of available large-scale GWAS datasets. Additionally, MR analysis excludes confounding effects by using SNPs associated with exposure as genetic instruments, which also reduces the adverse effects of inaccurate data on hindering identification of relationships between exposure and outcome.

The present MR analysis has several limitations. First, some subjects may have overlapped between the two data sets with respect to the estimates of instrument-exposure and instrument-outcome, which could lead to inflated type 1 error rates and false-positive findings (Burgess et al., 2016).

Furthermore, MR analyses are based on the GWAS. GWAS requires numerous subjects, often in multiple cohorts. Disease definition can differ among different cohorts. Third, we mostly included studies involving a predominantly European population with few individuals of other ancestries (mixed); hence, the present results may not be applicable to other racial backgrounds. Nevertheless, the present results support the results of previous epidemiology studies and promote further studies in this field.

## Data Availability

The summary statistics for T2D-related trait were obtained through large-scale genome-wide association (GWAS) meta-analyses of 133,010 non-diabetic individuals from the collaborating studies within the Meta-Analysis of Glucose and Insulin related traits Consortium (MAGIC) 

https://www.magicinvestigators.org/

## 6 Author Contributions

J.J. analyzed and interpreted the results and wrote the manuscript. S.W. and S.L. designed the study. All authors revised this paper critically for important intellectual content.

## 8 Acknowledgements

None

## 9 Data Availability Statement

All datasets presented in this study are included in the supplementary material and available at https://www.mrbase.org/, and https://gwas.mrcieu.ac.uk/.

